# Predicting the likelihood of elevated transaminases in lung cancer patients undergoing immune checkpoint blockade

**DOI:** 10.1101/2024.05.11.24307220

**Authors:** Dmitrii Shek, Bo Gao, Matteo S Carlino, Adnan Nagrial, Tania Moujaber, Deme Karikios, Ines Pires da Silva, Scott A Read, Golo Ahlenstiel

**Author notes:** <u>Corresponding author:</u> **Professor Golo Ahlenstiel**, Marcel Crescent, Blacktown, NSW, Australia; 2148, *E-mail:.

## Abstract

**Background:** The recent implementation of immune-checkpoint inhibitors (ICIs) has markedly changed the management and clinical outcomes for patients with advanced non-small cell lung cancer (NSCLC). Despite higher efficacy, ICIs are associated with a range of immune-related adverse events (irAEs). This retrospective study aimed to investigate the incidence of ICI-related transaminitis/hepatitis in NSCLC patients as well as to establish if any pre-treatment clinical parameters can predict the onset of liver toxicity.

**Methods:** We examined medical records of N=420 NSCLC patients treated across two health districts in Sydney, Australia between 2016 to 2020. R packages (corrplot and ggcorrplot) were used to construct correlation matrices and to calculate the correlation p-value using Spearman method. Logistic regression models were used to determine association of clinical parameters with elevated LFTs.

**Results:** N=185 patients were considered eligible. n=37 (20%) had elevation of liver transaminases at any stage post-ICI commencement, although only n=10 were deemed as those having ICI-related hepatitis. Most of these patients (n=29 [78%]) developed elevated LFTs within 3 months post-therapy initiation. Regression model established that pre-treatment ratio of serum protein (SP) to total bilirubin (TB) showed significant association with the elevated transaminases. Moreover, most of the patients (n=34 [94%]) with elevated LFTs had SP/TB<4. Using a second cohort of melanoma patients, the linear regression model did not establish a significant association between the SP/TB ratio and elevated LFTs. Nonetheless, n=31 (70.5%) patients with immune-mediated transaminitis in the melanoma cohort had SP/TB<4.

**Conclusion:** Our study has established a clinical risk factor associated with the elevation of LFTs in NSCLC patients, thus potentially enabling their prediction at the pre-treatment stage. However, its predictive accuracy was not confirmed in melanoma patients, stressing that ‘*one size does not fit it all*’ when developing predictive scores for irAEs in different patient cohorts.

## INTRODUCTION

Lung cancer is the leading cause of cancer-related mortality worldwide, resulting in over 1.5 million deaths every year ^1,2^. Smoking is the strongest risk factor, although more than 500,000 lung cancers are detected every year in patients who are lifetime non-smokers ^2,3^. The recent discovery of immune-checkpoint inhibitors (ICIs) ^4^ has markedly changed the treatment landscape for patients with advanced non-small cell lung cancer ^5^. ICIs are monoclonal antibodies (mAbs) ^6^ targeting immune checkpoint proteins including cytotoxic T-lymphocyte-associated antigen-4 (CTLA-4), programmed cell death-1 (PD-1) and programmed cell death ligand-1/2 (PD-L1/2) on cancer, T- and antigen-presenting cells ^7^.

The phase III Keynote-024 study reported that patients with advanced lung cancer, PD-L1 expression ≥50% and treated with anti-PD-1 mAb *Pembrolizumab* (PEMBRO) had superior overall survival (OS) of 30 months compared to 14.2 months in patients undertaking platinum doublet chemotherapy (hazard ratio [HR]: 0.63; 95% CI, 0.47 to 0.86; p=0.002) ^8^. Other studies that have evaluating the efficacy of *Nivolumab* (NIVO) in patients with advanced NSCLC as a second line approach have also established that OS and PFS rates were significantly higher in NIVO groups ^9^ compared to conventional chemotherapy ^10^. These results shifted the paradigm of advanced NSCLC treatment; PEMBRO is a current standard of care (SoC) for patients with PD-L1 expression ≥50% and NIVO for second line option in patients progressed on chemotherapy ^11^. For those who have PD-L1 expression <50%, PEMBRO and platinum doublet chemotherapy is a current first-line treatment ^11^.

Despite higher efficacy, ICIs are associated with a range of immune-related adverse events (irAEs) ^12^. ICI-mediated liver damage drives the elevation of liver function tests (LFTs) and occurs in up to 6% of patients treated with a PD-1 agent alone, and up to 30% treated with dual checkpoint blockade ^13^. Severe (> 5x upper limit normal [ULN]) liver toxicities are managed with cessation of treatment, administration of oral or intravenous steroids and/or other immunosuppressive drugs which can unfortunately result in cancer progression and reduced survival ^14^. Although there has been explicit research on irAEs in melanoma patients, as the first ICI was approved for melanoma, the understanding of irAEs in the context of immunotherapy for lung cancer is limited. This retrospective study was aimed to longitudinally monitor levels of liver transaminases in real-world cohort of patients with advanced NSCLC treated with ICIs. Secondly, we aimed to identify pre-treatment blood parameters that were associated with the onset of LFT elevation to develop a predictive score to identify patients susceptible to liver-specific irAEs.

## METHODS

### Study design

Research data collection and analysis was conducted as part of the NCT04595734 study, which has been approved by the Western Sydney Local Health District (WSLHD) Human Research Ethics Committee (ethics reference number: 2019/ETH13228) to be conducted at Blacktown Mt Druitt, Westmead and Nepean Hospitals. This study followed Strengthening the Reporting of Observational Studies in Epidemiology (STROBE) reporting guidelines for observational cohort studies.

### Patient cohort

We retrospectively collected clinical and pathological data from patients with advanced NSCLC treated at tertiary public hospitals across Western Sydney and Nepean Blue Mountains Local Health Districts in New South Wales, Australia. We examined medical records of eligible patients diagnosed from 2016 to 2020. Eligibility criteria included: (1) treatment with ICIs; (2) absence of liver metastases on baseline CT scans; (3) LFTs within the normal range (< 40 U/L) prior to ICI commencement; (4) absence of pre-existing hepatobiliary disorders and/or hepatic failure. Medical history and clinicopathological data were collected from electronic medical records. In addition, we used a de-identified cohort of n=140 melanoma patients from Melanoma Institute Australia treated with dual checkpoint blockade as a second cohort to elucidate the established risk factor in a different tumour.

### Clinical data collection

Examined data included: Eastern Cooperation Oncology Group (ECOG) performance status, smoking status, full blood count, electrolytes, urea, creatinine, LFTs, comorbidities, medications, mutations including PD-L1 status and other relevant details from medical history. In addition, we calculated the baseline AST to Platelet Ratio Index (APRI) ^15^ and Fibrosis-4 (FIB-4) scores ^16^ for each patient to determine the pre-existing liver fibrosis and its role for immunotherapy-related liver damage.

### Study objective

The goal of this study was to identify and enumerate the number of patients with elevated LFTs and to establish pre-treatment clinical parameters that were statistically associated with elevated liver transaminases. Medical records, particularly the blood test results were retrospectively examined from the day of ICI commencement to the last available test result or the data collection cut-off (December 2020), whichever occurred first.

### Statistical analysis

Statistical considerations were calculated using GraphPad Prism version 9.0 and R version 3.5.0 (R Foundation for Statistical Computing, Vienna, Austria) software. Variables were summarized using descriptive statistics. The non-parametric Mann-Whitney test was used to compare parameters among patients with and without irAEs. The non-parametric Spearman test was used to validate correlation of pre-treatment blood test parameters with elevated LFTs in our retrospective cohort. R packages (corrplot and ggcorrplot) were used to construct correlation matrices and to calculate the correlation p-value using Spearman method. Logistic regression models were used to determine association of clinical parameters with elevated LFTs. Results were considered significant with two-tailed p-value < 0.05. The study followed the Transparent Reporting of a multivariable prediction model for Individual Prognosis or Diagnosis guideline on reporting predictive models ^17^.

## RESULTS

### Clinical characteristics of the study cohort

We examined medical records of N=420 NSCLC patients treated across the two largest health areas in Sydney, Australia: (1) Western Sydney district (Blacktown Mt Druitt and Westmead Hospitals), (2) Nepean and Blue Mountains district (Nepean Hospital). Among them, N=185 patients were considered eligible, and their medical records were retrospectively examined (Figure 1). The median cohort age was 71 years old and n=97 (52.4%) identified themselves as male. PD-L1 expression > 50% was present in n=44 (23.8%). The top-3 comorbidities in the study cohort were ischemic heart disease, arterial hypertension, chronic obstructive pulmonary disease. Table 1 summarises the demographic and clinical features of the study cohort.

**Figure 1.**
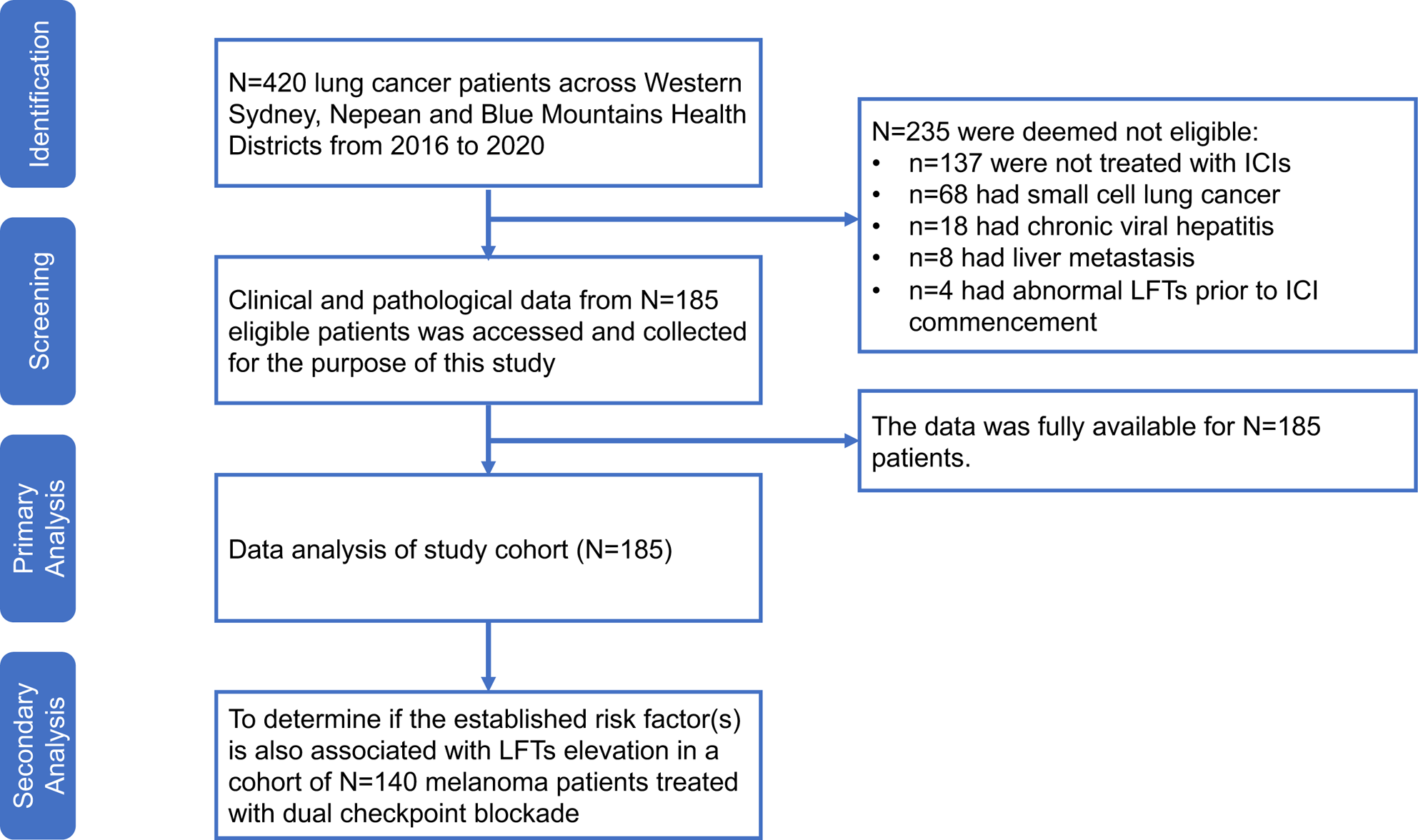
STROBE flow diagram of examined patients and their medical records. The diagram represents the workflow of this retrospective cohort study. Abbreviations: LFTs – liver functional test; ICI – immune-checkpoint inhibitor.

**Table 1.** Clinical summary of the study cohort. ECOG – eastern cooperation oncology group; ICI – immune-checkpoint inhibitor; COPD – chronic obstructive pulmonary diseases; BA – bronchial asthma; HTN – hypertension; IHD – ischemic heart diseases; MDD – major depressive disorder; GORD – gastroesophageal reflux diseases, T2DM – type 2 diabetes mellitus, ULN – upper limit normal (40 U/l). Prior smoking status was defined as a person who stopped smoking at least 2 years prior to treatment commencement.

| Characteristic | Single ICI, n | Dual ICI, n | ICI + Chemo, n | Total, n (%) |
| --- | --- | --- | --- | --- |
| *N* | 58 (31%) | 55 (30%) | 72 (39%) | 185 (100%) |
| *Ethnicity:* |  |  |  |  |
| Caucasian | 43 | 33 | 47 | 123 (67%) |
| Asian | 12 | 17 | 12 | 41 (22%) |
| African | 2 | 2 | 6 | 10 (5%) |
| Indigenous Australian | 1 | 3 | 7 | 11 (6%) |
| *ECOG performance status at diagnosis:* |  |  |  |  |
| 0 | 45 | 41 | 51 | 137 (74%) |
| 1 | 11 | 9 | 14 | 34 (18%) |
| 2 | 2 | 5 | 7 | 14 (8%) |
| *Histology:* |  |  |  |  |
| Adenocarcinoma | 50 | 48 | 49 | 147 (80%) |
| Squamous cell carcinoma | 7 | 5 | 18 | 30 (16%) |
| Large cell carcinoma | 1 | 2 | 5 | 8 (4%) |
| *Line of ICI:* |  |  |  |  |
| First line | 22 | 14 | 10 | 46 (25%) |
| Second line | 35 | 38 | 51 | 124 (67%) |
| Beyond second line | 1 | 3 | 11 | 15 (8%) |
| *Smoking status:* |  |  |  |  |
| Current | 12 | 10 | 22 | 44 (24%) |
| Prior | 28 | 30 | 44 | 102 (55%) |
| Never | 18 | 15 | 6 | 39 (21%) |
| *LFTs elevation:* |  |  |  |  |
| < 3 x ULN | 6 | 7 | 14 | 27 (15%) |
| > 3 x ULN | 4 | 5 | 1 | 10 (5.4%) |

Within the cohort of 185 patients, n=37 (20%) had elevated liver transaminases at any time point post-ICI commencement. Among these patients, n=8 were treated with a single anti-PD-1 agent, n=2 with a single anti-PD-L1 agent, n=12 with dual CTLA-4/PD-1 blockade and n=15 with a combination of platinum-based chemotherapy + ICI agent. Most of the patients (n=29 [78%]) developed elevated LFTs within 3 months post-therapy initiation. The rest of the patients (n=8 [22%]) developed late-onset (> 3 months post cycle 1) transaminitis. Interestingly, the majority (n=27) of patients with low grade LFT elevation had no clinical symptoms and were not treated with steroids or other immunosuppressive agents. Among n=10 patients (5.4% from the total study cohort), LFT elevation (grade 2 ^13^ and higher) was classified as ICI-related hepatitis. These patients ceased immunotherapy and received oral prednisolone (n=7) or intravenous dexamethasone followed by oral corticosteroids (n=3). Patients with high-grade ICI-related hepatitis were treated either with single anti-PD-1 agent (n=4), dual CTLA-4/PD-1 blockade (n=5) and combination of platinum-based chemotherapy + ICI agent (n=1). All patients developed ICI-related hepatitis between weeks 8-10 post-ICI commencement. N=6 patients with ICI-related hepatitis were eventually rechallenged with single anti-PD-1 agent. N=4 patients with grade 3 hepatitis passed away within the 2 months following ICI cessation. No patients with elevated transaminases were treated with the anti-CTLA-4 drug *Ipilimumab* (IPI) at a dosage of 3 mg/kg ^18^.

### Pre-treatment factors associated with elevation of liver transaminases

We next sought to identify any pre-treatment clinical or demographic parameters that are associated with the ICI-mediated elevation of liver transaminases. First, we aimed to determine if previously proposed markers of ICI-related toxicity, such as BMI ^19^ or neutrophil-to-lymphocyte ratio (NLR) ^20^ are correlated with the elevated transaminases in our study cohort. The mean BMI and NLR values among patients (1) without and (2) with elevated LFTs were found to be: (1) 26.4 (standard deviation [SD] = 1.2) and 7.4 (SD = 0.91), (2) 25.6 (SD = 3.1) and 6.78 (SD = 1.03) respectively. Analysis using Student’s t-test revealed no significant difference of the examined parameters. Linear regression did not demonstrate an association between pre-treatment BMI (Figure 2A) or NLR (Figure 2B) and elevated levels of ALT and/or AST at the toxicity onset. Furthermore, no correlation was established between pre-treatment BMI and NLR and the level of liver transaminases in patients without immune-related transaminitis/hepatitis.

**Figure 2.**
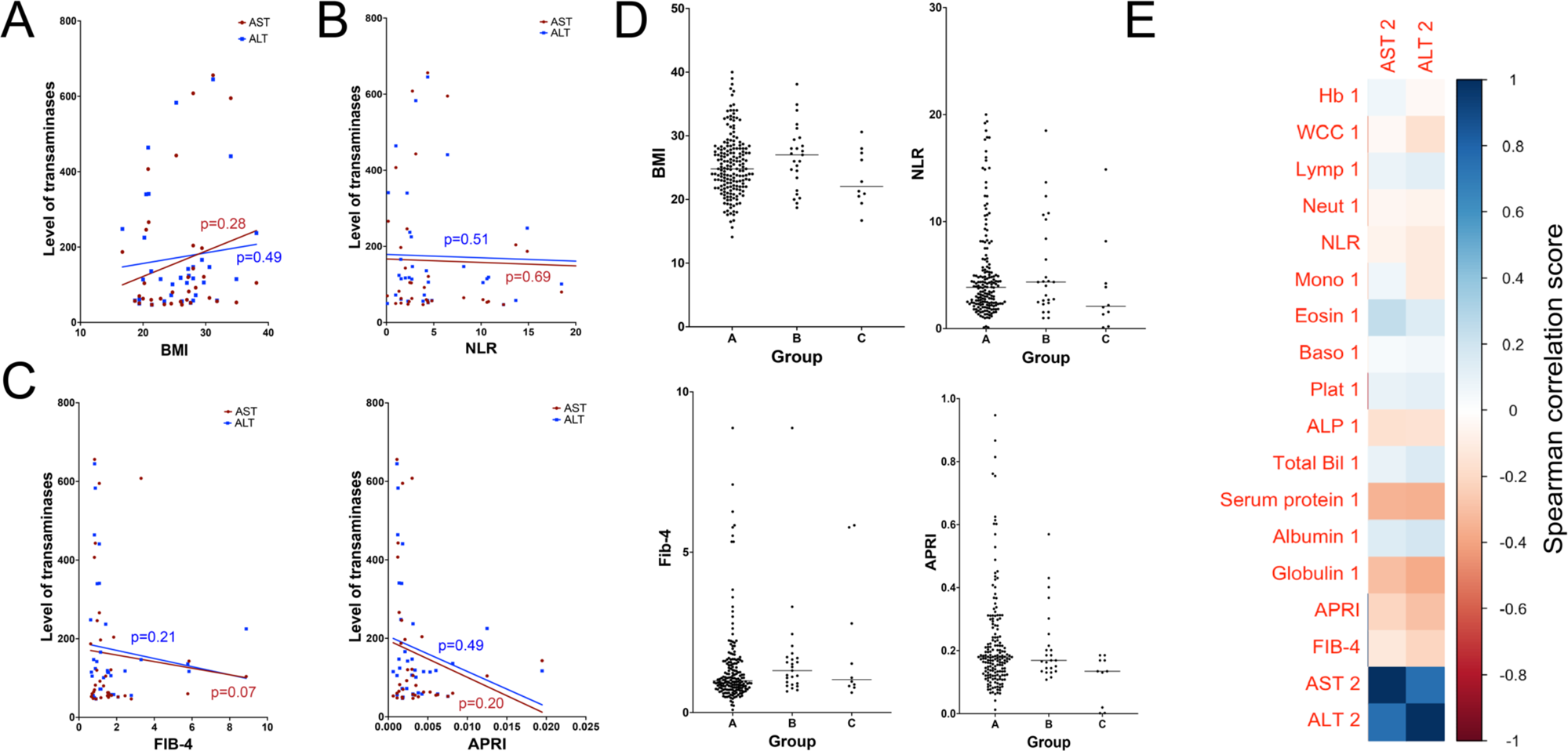
Association of pre-treatment clinical parameters and LFTs elevation. No statistical significance was established between any pre-treatment clinical parameters and APRI, FIB-4, BMI, NLR scores with the elevated ALT and AST. **A, B, C** – linear regression model determining the association between BMI, NLR, fibrosis scores and elevated transaminases. **D** – Selected parameters in the study cohort (A – patient with no LFTs elevation, B – patients with LFTs elevation but not deemed as those having ICI-related liver injury, C – patients with diagnosed ICI-related hepatitis). **E** – Spearman correlation matrix elucidating the correlation of pre-treatment blood test parameters with elevated LFTs.

Next, we examined the relationship between pre-treatment fibrosis scores including the AST to platelet ratio index (APRI) and (2) Fibrosis-4 (FIB-4) scores with elevated LFTs. The mean APRI and FIB-4 scores among (1) without and (2) with elevated LFTs were found to be: (1) 0.091 (SD = 0.0023) and 1.83 (SD = 0.19), (2) 0.046 (SD = 0.0019) and 1.78 (SD = 0.15) respectively (p>0.05). In addition, no significant association between pre-treatment fibrosis scores and elevated transaminases was established (Figure 2C, 2D). Lastly, a Spearman correlation matrix (Figure 2E) was generated to examine the association between any pre-treatment blood test results and transaminase values, demonstrating no significant relationships.

We next sought to determine whether pre-treatment LFT ratios (*e.g*., AST/ALT) may predict ICI-mediated liver toxicity in our cohort. We first generated all possible ratios from a set of liver-related clinical parameters (globulin, albumin, serum protein, total bilirubin, gamma-glutamyl transferase [GGT], alkaline phosphatase [ALP]) using R. Next, we used a Spearman correlation matrix to determine if any of newly established ratios are associated with ICI-mediated liver toxicity (ALT and AST). Interestingly, our results identified a subset of LFT ratios that significantly correlated with elevated AST and ALT during acute liver toxicity (Figure 3A). To further determine their predictive ability, we used a multiple regression model to determine if the established parameters can indeed predict the elevation of LFTs. Interestingly, only ratio of serum protein (SP) to total bilirubin (TB) showed significant association with both elevated AST and ALT (Figure 3B). To further determine the role of SP/TB ratio in predicting the elevation of LFTs we calculated the value of SP/TB ratio across all examined patients. Notably, most of the patients (n=34/37 [94%]) with elevated LFTs had SP/TB < 4. In contrast among patients with normal level of transaminases only n=42/148 (28%) had the ratio less than 4. The predictive value of the proposed score is summarised on Figure 3C, demonstrating a 44.7% positive predictive value (PPV) and a 97.2% negative predictive value (NPV). Upon examining the individual LFT values in both groups, the mean SP was 62.7 g/L and 70.5 g/L in patients with and without elevated LFTs, respectively (p>0.05) and the mean TB was 17.2 µmol/L and 10.8 µmol/L in patients with and without transaminitis, respectively (p>0.05).

**Figure 3.**
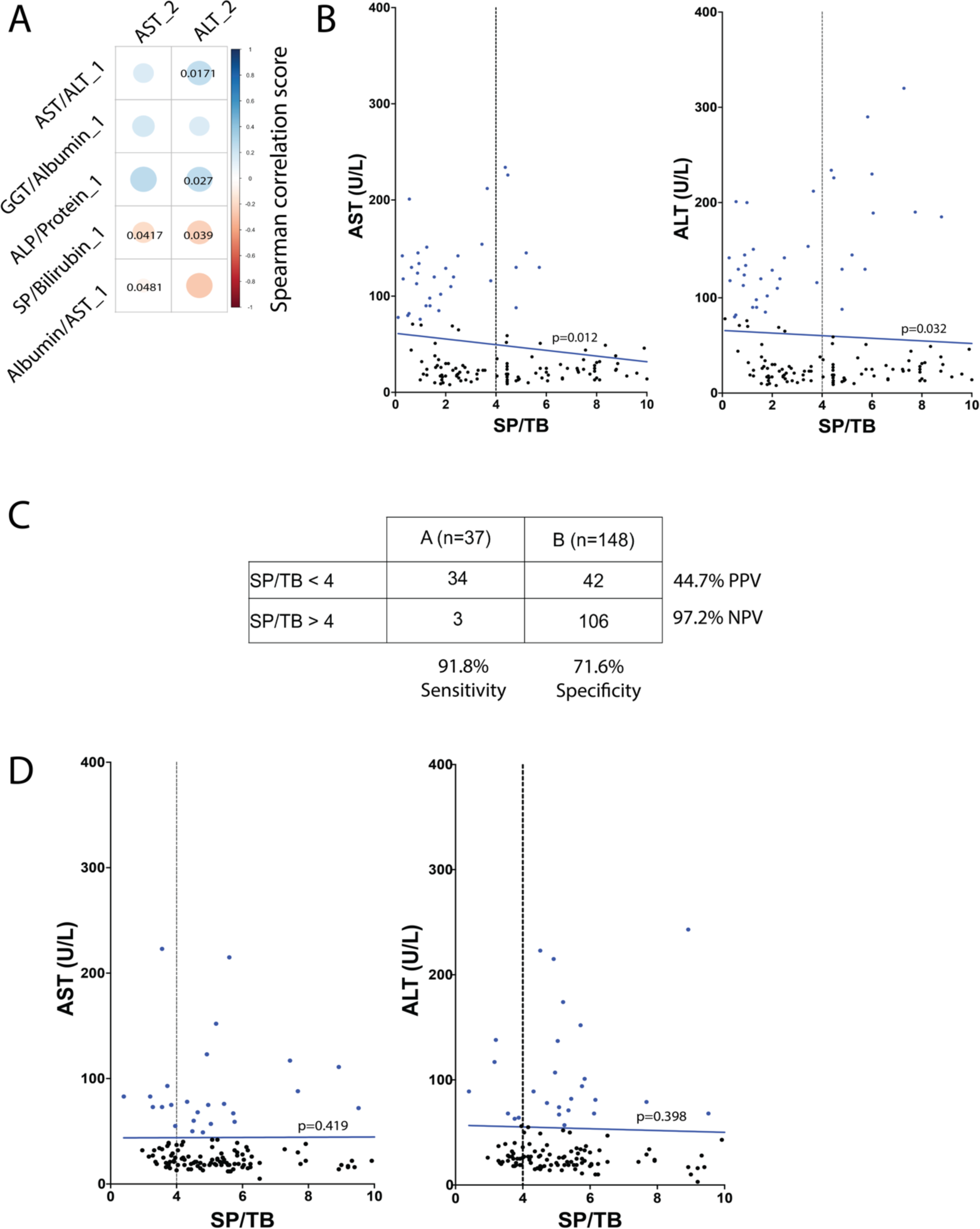
Clinical factors associated with the elevated transaminases. **A** – pre-clinical ratios significantly correlated with elevated LFTs. Spearman’s rank correlation method was used, due to assumption of non-parametric data distribution. **B** – Linear regression model established a significant association between pre-treatment SP/TB ratio and elevated ALT and AST in the examined cohort of NSCLC patients. **C** – Predictive value of the ratio SP/TB in the cohort of NSCLC patients with elevated LFTs. **D** – Linear regression model established no significant association between pre-treatment SP/TB ratio and elevated LFTs in the validation cohort of melanoma patients. Group A (blue) are patients who had elevated LFTs post-immunotherapy and group B (red) those who did not develop any hepatic side effects. Abbreviations: SP serum protein; TB – total bilirubin; PPV – positive predictive value; NPV – negative predictive value.

### Assessing the predictive accuracy of the SP/TB ratio in a melanoma cohort

To further elucidate the established risk factor, we retrospectively examined a second cohort of N=140 melanoma patients from Melanoma Institute Australia treated with dual CTLA-4/PD-1 blockade. N=38 (27%) received anti-CTLA-4 drug IPI in dosage of 1 mg/kg, while the rest of the cohort (n=102 [73%]) received 3 mg/kg. Within this cohort n=44 (31%) developed ICI-related hepatitis. Among them n=4 (9%) had grade 1 (no steroid treatment received), n=16 (36%) had grade 2 (treated with oral prednisolone) while n=24 (55%) developed high-grade toxicity (grade 3 and higher) who were treated with multiple immunosuppressive agents including corticosteroids and/or mycophenolate mofetil. Only n=3 (12.5%) patients who developed ICI-related hepatitis of grade 3 and higher were treated with IPI 1 mg/kg, while the rest (n=21 [87.5%]) were treated with IPI 3 mg/kg. We collected the blood test results data from pre-treatment and after 2 cycles of immunotherapy to validate if the established pre-treatment ratio SP/TB would be also significantly associated with the elevation of LFTs. Interestingly the linear regression model did not establish a significant association of SP/TB ratio with the elevated LFTs (Figure 3D). To ascertain whether there was a significant difference in baseline LFTs that may influence the predictive efficacy of the SP/TB ratio, we compared both study and melanoma cohorts. It is worth mentioning that no significant differences in pre-treatment levels of ALT or AST were established between two examined cohorts (Table 2, supplementary table 1). Nonetheless, we established a significant difference in the level of total protein, SP/TB, GGT/albumin, and albumin/AST ratios between the examined cohorts, although its significance is not yet understood. Finally, n=31 (70.5%) patients with immune-mediated transaminitis in the melanoma cohort had SP/TB < 4 further stressing potential role of SP/TB ratio in predicting the evolving elevation of liver transaminases.

**Table 2.**
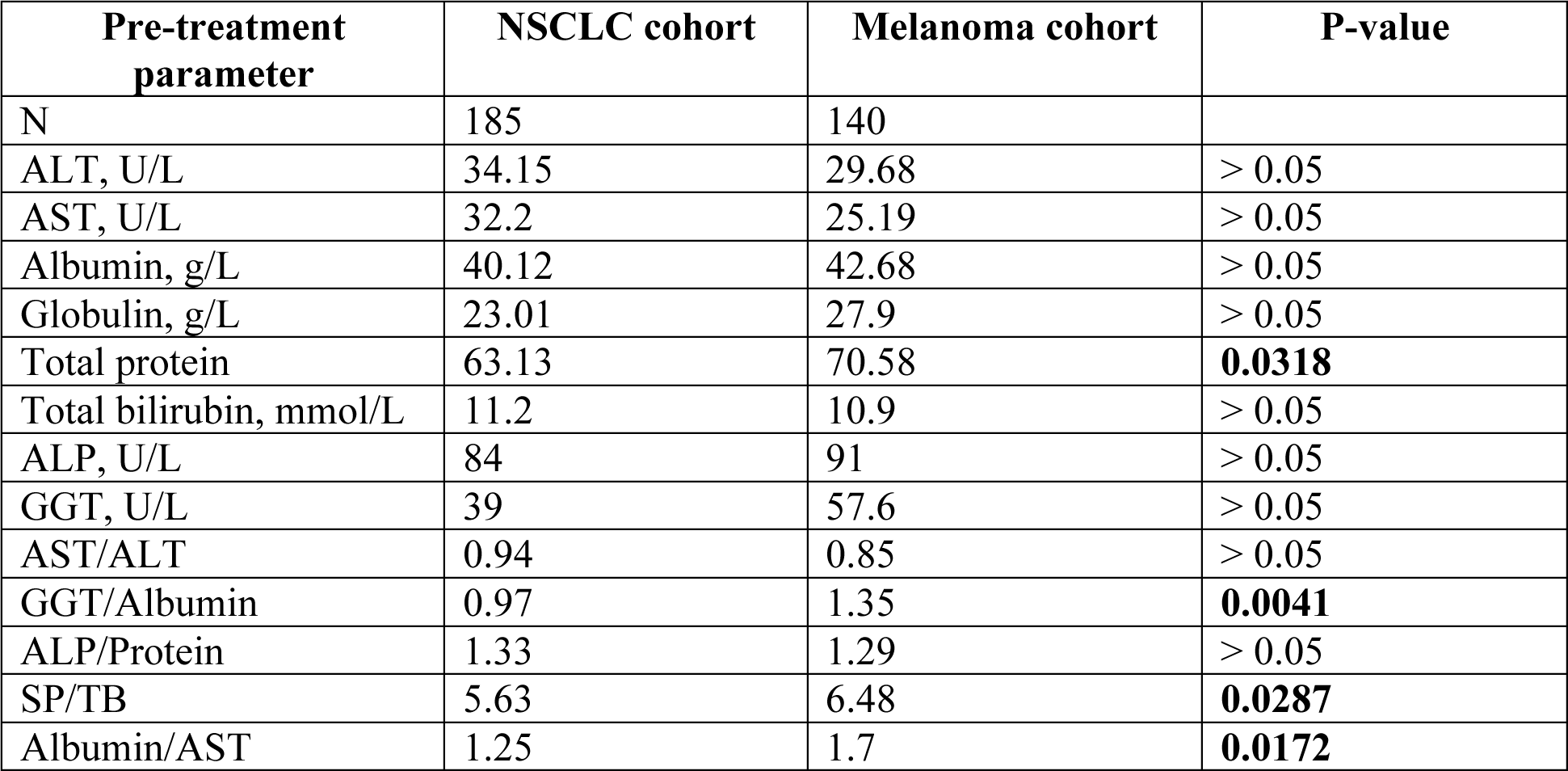
Mean pre-treatment liver function test results and ratios which were significantly associated with the elevated transaminases in study cohort and compared to second (melanoma) cohorts. P-value was calculated using two-tailed t-test and <0.05 was considered as significant.

## DISCUSSION

Immunotherapy has shifted the therapeutic landscape for multiple solid malignancies ^21^. By targeting inhibitory checkpoint receptors expressed on both T and antigen-presenting cells, these drugs can boost proliferation of cytotoxic CD8+ T cells thus retarding cancer progression ^21^. Unfortunately, the abundant activation of effector T cells is commonly associated with adverse events for which the aetiology is not fully understood ^21^. In this study we established that 20% of NSCLC patients treated with ICIs experienced elevation of liver transaminases with the majority within the first 3 months post-ICI commencement. Interestingly, only 10/185 (5.4%) of cases developed LFT elevation >3 x ULN, resulting in a diagnosis of immunotherapy-related hepatitis. Through our analysis we established that the ratio of SP/TB was statistically associated with the elevated LFTs in this cohort. A SP/TB cut-off of four was introduced to stratify patients based on their likelihood of developing elevated LFTs.

To elucidate the physiological basis of this scoring system and its relevance in predicting hepatic irAEs, it is crucial to stress that liver is a distinctive immunological organ situated at the intersection of systemic and intestinal circulation ^22^. The liver possesses the capability to adhere activated T cells (both CD4+ and CD8+) through the interaction of T cell α4β1-integrin and lymphocyte function-associated antigen 1 (LFA-1) with sinusoidal endothelial cell or Kupffer cells vascular cell adhesion molecule 1 (VCAM-1) and intercellular adhesion molecule 1 (ICAM-1) ^23^. This retention of T cells within the liver results in their interaction with Kupffer cells via Fas binding ^6,24^, triggering the production of tumour necrosis factor-alpha (TNF), ultimately inducing CD8+ T cell mediated hepatocyte apoptosis ^25,26^. Hepatocyte injury is likely to be exacerbated during ICI therapy due to the inhibition of natural barriers that regulate T cell activation. The subsequent destruction of hepatocytes leads to compromised synthetic liver function, characterized by reduced protein synthesis including albumin ^27^, representing >50% of serum protein ^28^, the numerator in the SP/TB ratio. Simultaneously, hepatocyte damage impairs the uptake and conjugation of indirect bilirubin, resulting in an increase of both indirect and total bilirubin levels in blood ^29^, consequently increasing the denominator and diminishing the SP/TB ratio. This may explain why patients with elevated LFTs in our study exhibited a lower SP/TB ratio compared to those without elevated transaminases. While a significant mean difference in individual values of SP and TB between the examined groups (with and without ICI-mediated transaminitis) of patients was not established, the mean SP/TB ratio in patients with elevated LFTs was lower at 3.6 compared to 6.5 in patients without elevated transaminases.

Interestingly, the SP/TB ratio did not demonstrate any predictive accuracy in the cohort of melanoma patients, although n=31 (70.5%) patients with immune-mediated transaminitis had a SP/TB ratio < 4. While tumours may be treated with analogous ICI-based regimens, disparities in their histological architecture, mutational burden and immune microenvironment can impact the predictive capacity of established risk factors ^30^. While numerous studies ^31^ have focused on the identification of pan-cancer genomic, proteomic and clinical markers for predicting irAEs, it is crucial to recognise that different tumours have variable intrinsic resistance to ICI therapy leading to a variable efficacy and safety outcomes^32^. Moreover, regardless of tumor type or therapeutic regimen, the incidence of irAEs is also influenced by patient-specific characteristics such as age, gender, comorbidities and concurrent medications ^33^. Furthermore, the possibility of irAEs to manifest either early or late following commencement of ICI therapy, suggests that additional predisposing factors have not yet been identified ^33^. In the future, it may therefore be necessary to define predicitve markers tailored to each unique cancer type, therapeutic regimen, patient cohort and combinations thereof.

Despite the differences observed between two cohorts, this study has provided novel data on potential clinical predictive factor associated with the elevation of LFTs in lung cancer patients. There are, however, some limitations worth mentioning. Firstly, this is a retrospective study and a subject to selection bias due to incompletely or inaccurately recorded medical information. Secondly, it is possible that unidentified confounding factors related to this study population have contributed to differences in liver toxicity identified in this study. Thirdly, we decided to empirically explore the correlation between different pre-treatment ratios of LFTs and ICI-related transaminitis/hepatitis, without any clear rationale preceding our decision.

To sum up, this study has provided a comprehensive overview of liver injury in lung cancer patients treated with ICIs. Moreover, we have established a clinical risk factor associated with the evolving elevation of LFTs, thus potentially enabling the prediction at pre-treatment stage. While further validation through larger prospective studies is warranted, the observed association provides a valuable starting point for developing more specific monitoring strategies and personalized interventions to mitigate the risk of hepatotoxicity in the era of immunotherapeutic advancements. Future establishment of clinical and biological markers specific to each cancer type will markedly benefit patient outcomes by improving safety parameters of immunotherapy.

## Data Availability

All data produced in the present study are available upon reasonable request to the authors

## Acknowledgements

We want to acknowledge (1) Amy Phu from Westmead Hospital for assisting with the database maintenance and (2) Jodie McMurray for helping with clinical data collection at Nepean Hospital.

## Author contributions

Dmitrii Shek, Bo Gao, Scott Read, Golo Ahlenstiel created the concept and design of the study. Matteo S Carlino, Bo Gao, Adnan Nagrial, Tania Moujaber, Ines Pires da Silva, and Deme Karikios provided with study materials. Dmitrii Shek, Golo Ahlenstiel conducted the data collection and assembly. Dmitrii Shek, Scott Read and Golo Ahlenstiel performed statistical analysis and interpretation. Golo Ahlenstiel provided administrative support. Dmitrii Shek, Scott Read, Golo Ahlenstiel wrote the manuscript and all authors equally contributed to editing and reviewing this manuscript prior to submission.

## Ethics approval

This retrospective study was conducted in accordance with the National Health and Medical Research Council’s National Statement on Ethical Conduct in Human Research and the CPMP/ICH Note for Guidance on Good Clinical Practice. The study has been approved by Western Sydney Local Health District (WSLHD) Human Research Ethics Committee (HREC) with Research Office File Number 6258 (2019/ETH13228).

## SUPPLEMENTARY DATA

**Table 1.** Additional clinical and demographic parameters from lung cancer and melanoma cohorts. P-value was calculated using two-tailed t-test and <0.05 was considered as significant.

| Parameter | NSCLC cohort | Melanoma cohort | P-value |
| --- | --- | --- | --- |
| Median age | 71 (56 to 82) | 61 (43 to 74) | **0.0281** |
| G1-2 hepatitis, n (%) | 27 (15%) | 20 (14.3%) | > 0.05 |
| G3-4 hepatitis, n (%) | 10 (5.4%) | 24 (17%) | **0.001** |
| Median (IQR) number of weeks for hepatitis onset | 12 (8 to 25) | 13 (10 to 32) | > 0.05 |
| Haemoglobin (mean, g/L) | 152 | 138 | > 0.05 |
| White blood cells (mean, x10<sup>9</sup>/L) | 10.4 | 11.5 | > 0.05 |
| Lymphocytes (mean, x10<sup>9</sup>/L) | 1.57 | 2.27 | > 0.05 |
| Neutrophils (mean, x10<sup>9</sup>/L) | 7 | 8.35 | > 0.05 |
| Monocytes (mean, x10<sup>9</sup>/L) | 0.89 | 0.74 | > 0.05 |
| Eosinophils (mean, x10<sup>9</sup>/L) | 0.36 | 0.51 | > 0.05 |
| Basophils (mean, x10<sup>9</sup>/L) | 0.06 | 0.1 | > 0.05 |
| Platelets (mean, x10<sup>9</sup>/L) | 320 | 288 | > 0.05 |
| LDH (mean, U/L) | 213 | 251 | > 0.05 |

